# Accounting for intensity variation in image analysis of large-scale multiplexed clinical trial datasets

**DOI:** 10.1101/2023.05.19.23290216

**Authors:** Anja L Frei, Anthony McGuigan, Ritik RAK Sinha, Mark A Glaire, Faiz Jabbar, Luciana Gneo, Tijana Tomasevic, Andrea Harkin, Tim J Iveson, Mark Saunders, Karin Oein, Noori Maka, Francesco Pezella, Leticia Campo, Jennifer Hay, Joanne Edwards, Owen Sansom, Caroline Kelly, Ian Tomlinson, Wanja Kildal, Rachel S Kerr, David J Kerr, Håvard E Danielsen, Enric Domingo, TransSCOT consortium, David N Church, Viktor H Koelzer

## Abstract

Multiplex immunofluorescence (mIF) imaging can provide comprehensive quantitative and spatial information for multiple immune markers for tumour immunoprofiling. However, application at scale to clinical trial samples sourced from multiple institutions is challenging due to pre-analytical heterogeneity. This study reports an analytical approach to the largest multiparameter immunoprofiling study of clinical trial samples to date. We analysed 12,592 tissue microarray (TMA) spots from 3,545 colorectal cancers (CRC) sourced from more than 240 institutions in two clinical trials (QUASAR 2 and SCOT) stained for CD4, CD8, CD20, CD68, FoxP3, pan-cytokeratin and DAPI by mIF. TMA slides were multi-spectrally imaged and analysed by cell-based and pixel-based marker analysis. We developed an adaptive thresholding method to account for inter- and intra-slide intensity variation in TMA analysis. Applying this method effectively ameliorated inter- and intra-slide intensity variation improving the image analysis results compared to methods using a single global threshold. Correlation of CD8 data derived by our mIF analysis approach with single-plex chromogenic immunohistochemistry (IHC) CD8 data derived from subsequent sections indicates the validity of our method (Spearman’s rank correlation coefficients ρ between 0.63 and 0.66, p-value ≪ 0.01) as compared to current gold standard analysis approach. Evaluation of correlation between cell-based and pixel-based analysis results confirms equivalency (ρ > 0.8, p ≪ 0.01, except for CD20 in epithelium region) of both analytical approaches. These data suggests that our adaptive thresholding approach can enable analysis of mIF-stained clinical trial TMA datasets by digital pathology at scale for precision immunoprofiling.

## INTRODUCTION

Immunoprofiling, the assessment of the density, state and spatial distribution of immune cells, is a crucial part of the examination of a tumour and its microenvironment [1]. Immunoprofiling can help to identify predictive markers for better assignment of patients to treatment with immune modulators such as immune checkpoint inhibitors [2,3]. Considering the potential for severe adverse effects of these therapies, improved patient stratification is an urgent need [4]. At a more fundamental level, immunoprofiling can also provide new insights into cancer biology and contribute towards a better understanding of tumour progression. Multiplex immunofluorescence (mIF) imaging is a powerful method for spatially visualising multiple biomarkers at a cell-level resolution on a single slide [5], [6], enabling comprehensive cell phenotyping in the cancer microenvironment as compared to standard single-plex immunohistochemistry (IHC) staining. However, immunofluorescence (IF) imaging is prone to imaging artefacts when applied to clinical samples [7]. These artefacts can be caused by pre-analytical variation introduced by samples from multiple institutions, differences in fixation, embedding or by the imaging process [8]. During imaging, different types of fluorophores, exposure time, illumination intensity and bleaching effects can lead to variations in the resulting images [9]. Additionally, tissue-intrinsic fluorescence can distort the signal. In mIF imaging, channel cross-talk can add further complexity due to spectral overlap, which can be exacerbated when large panels are used due to the proximity of the different channels in the wave spectrum. Additionally, mIF staining and imaging technologies are cutting-edge technologies and, while single platforms themselves are standardised, no overarching standards across platforms exist. Therefore, considerable pre-analytical heterogeneity due to both staining and imaging of the histological slides can be frequently observed and the expected range of variation observed increases with the size and sample heterogeneity of the clinical cohorts under study. Tissue microarrays (TMAs) are a key tool for efficient analysis of large clinical trial cohorts [10] and allow simultaneous analysis of hundreds of patient samples on a single slide. TMA design including multiple punches from the same sample helps to capture intra-patient heterogeneity [11], making the downstream analysis more robust. Combining TMA technology with digital image analysis is an excellent approach to extract information from digitised TMA slides in a semi-automated manner [12]. Nevertheless, image analysis often relies on the assumption of relative homogeneity across the entire cohort which may not hold true for large multiplexed cohorts with samples from multi-centric clinical studies. Consensus approaches for quantitative image analysis in clinical cohorts are therefore of increasing importance as recognised by the consensus statement of the Society for Immunotherapy of Cancer (SITC) on best practices for multiplex IHC and IF staining and validation [13]. One method to handle signal variation by digital image analysis is pre-processing with the aim to normalise signal intensity and reduce signal variation within the dataset [14,15]. Ideally, normalisation reduces the impact of confounding pre-analytical factors while preserving biologically relevant heterogeneity. In the context of image analyses relying on thresholds, *adaptive thresholding* [16] can be applied to handle variation within a dataset instead of normalising the data beforehand. Adaptive thresholding denotes methods not using a single threshold for an entire dataset (*global threshold*) but choosing different thresholds (*local thresholds*) for different regions of analysis based on certain properties in the region to be analysed and its environment, thereby better reflecting intra-sample (e.g., at the pixel level in a single image) and inter-sample variation (e.g., at the image level in a cohort with multiple images) introduced by staining and imaging heterogeneity.

In this study, we develop a spatially resolved protocol for the detection and quantification of immune cells and systematically address different issues in the application of image analysis to multiplexed staining and imaging in application to the currently largest mIF clinical trial dataset reported in the literature. We report strategies for adaptive thresholding in the TMA setting when staining intensity varies substantially between and within images and systematically compare different strategies for cell-level quantification using both traditional cell segmentation techniques as well as pixel-based quantification metrics for individual channels. Last, we test the consistency and methodological robustness of our approach by comparison of the multiplexed data to the current gold standard of single-plex chromogenic IHC staining. The current study thus provides valuable data on the challenges and possible solutions for the quantitative image analysis of mIF data from clinical trials carried out in a series of institutions and multiple countries.

## MATERIALS AND METHODS

### Cohorts

The cohorts under study consist of high-risk stage II and stage III CRC cases from two clinical trials: QUASAR 2 (Q2) [17] & SCOT [18]. Q2 investigated whether the addition of bevacizumab to capecitabine improves the three-year disease-free survival after surgery of histologically proven stage III or high-risk stage II CRC and included 1952 patients from 170 hospitals in seven countries. The SCOT trial investigated whether three months of oxaliplatin-containing adjuvant chemotherapy is non-inferior to six months of the same treatment for high-risk stage II and stage III CRC. The SCOT trial included 6,088 patients from 244 centres in six countries: United Kingdom (England, Scotland, Wales and Northern Ireland), Denmark, Spain, Sweden, Australia and New Zealand. The CRC tissues from both trials were arranged into 79 TMA slides, containing 15,121 spots from 3,545 patients (between two and eight spots per patient; spot diameter 1.0 mm for Q2 and 0.6 mm for SCOT), see **Table 1**. SCOT tissue samples were processed at the NHS Greater Glasgow and Clyde. All TMA slides were stained with a Vectra Polaris Opal™ (Akoya Biosciences, Marlborough, MA, US) 7-plex IF panel (see **Table 2**) at the Translational Histopathology Laboratory, Department of Oncology, University of Oxford, United Kingdom. The multi-IF slides were processed by multispectral imaging on the Vectra Polaris (Akoya Biosciences, Marlborough, MA, USA) quantitative pathology imaging system at 20x magnification, spectrally unmixed using inForm (Akoya Biosciences, Marlborough, MA, USA) and stitched together using the HALO Image Analysis Platform (Indica Labs, Inc., Albuquerque, NM, USA), resulting in multi-channel IF whole-slide images (WSI) with a resolution of 0.4976 μm per pixel. See **Figure 1** for an example of a mIF image from the dataset and see **Figure 2** for visualisations of the variation observed in the image dataset.

**Table 1:** Dataset characteristics.

|  | Q2 | SCOT | Total |
| --- | --- | --- | --- |
| Number of TMA slides | 29 | 50 | 79 |
| Spot diameter | 1.0 mm | 0.6 mm | — |
| Number of spots | 3,465 | 11,656 | 15,121 |
| Number of valid spots | 2,650 | 9,942 | 12,592 |
| Number of cases | 1,195 | 2,350 | 3,545 |
| Number of cases with valid spots | 1,120 | 2,350 | 3,470 |
| Number of spots per case | 2 (120 cases) or 3 (1,075 cases) | 4 (1,786 cases) or 8 (564 cases) | — |
| Amount of analysed area | 2,624.66 mm<sup>2</sup> | 3,310.78 mm<sup>2</sup> | 5,935.44 mm<sup>2</sup> |
| Number of classified cells | 17,886,688 | — | — |

**Table 2:**
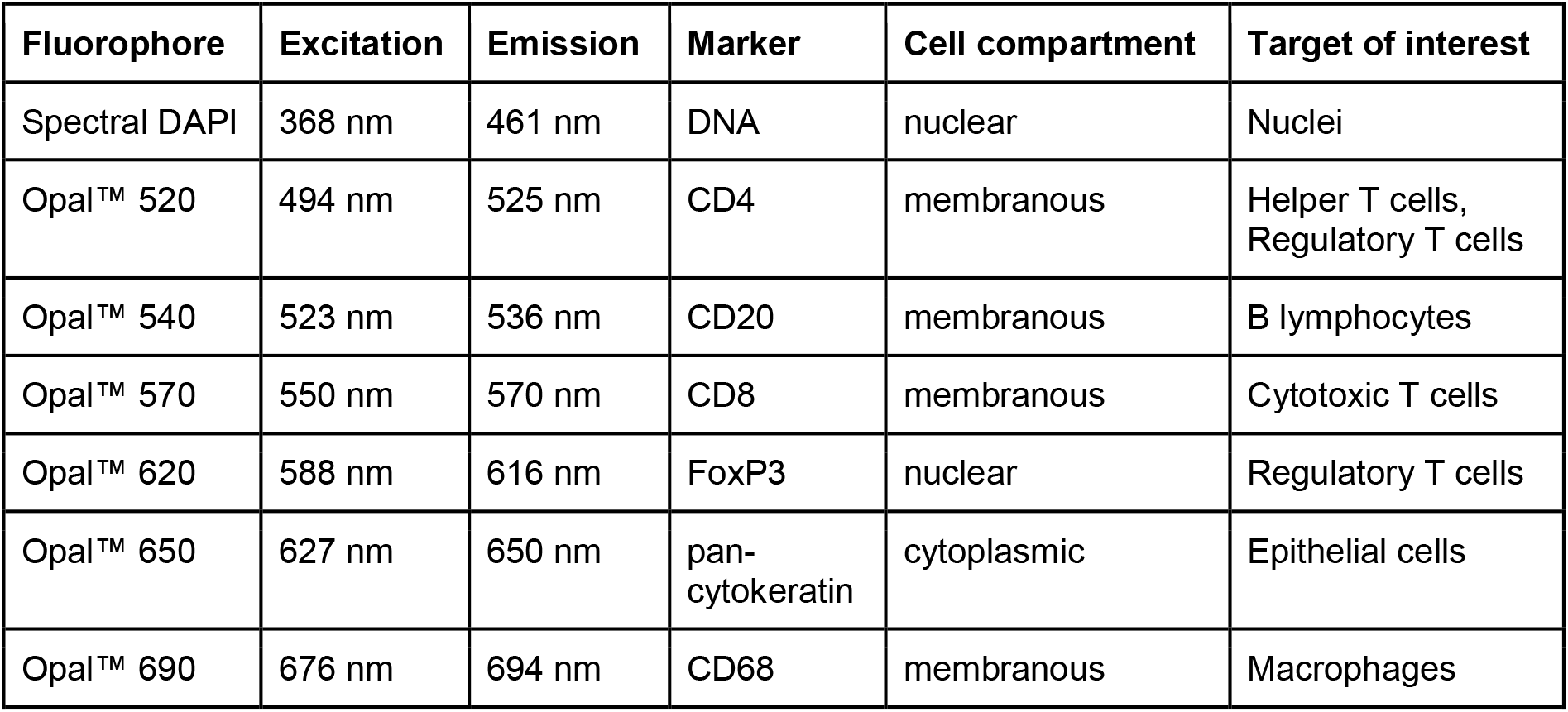
Marker panel.

**Figure 1:**
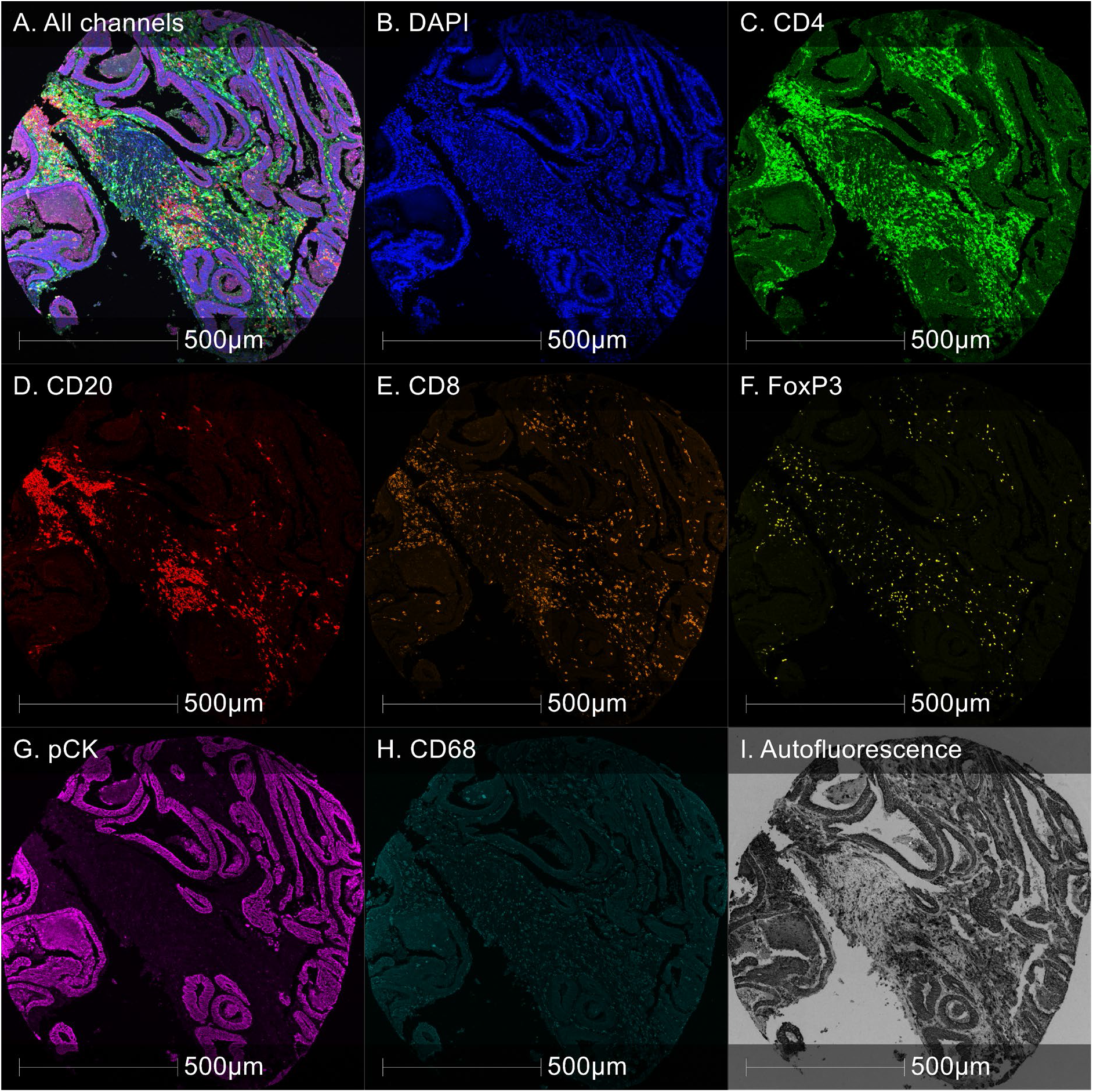
Example of 7-plex colorectal cancer TMA spot image from Q2 cohort. **A**. All channels combined. **B-H**. Individual marker channels (pCK: pan-cytokeratin). **I**. Autofluorescence.

**Figure 2:**
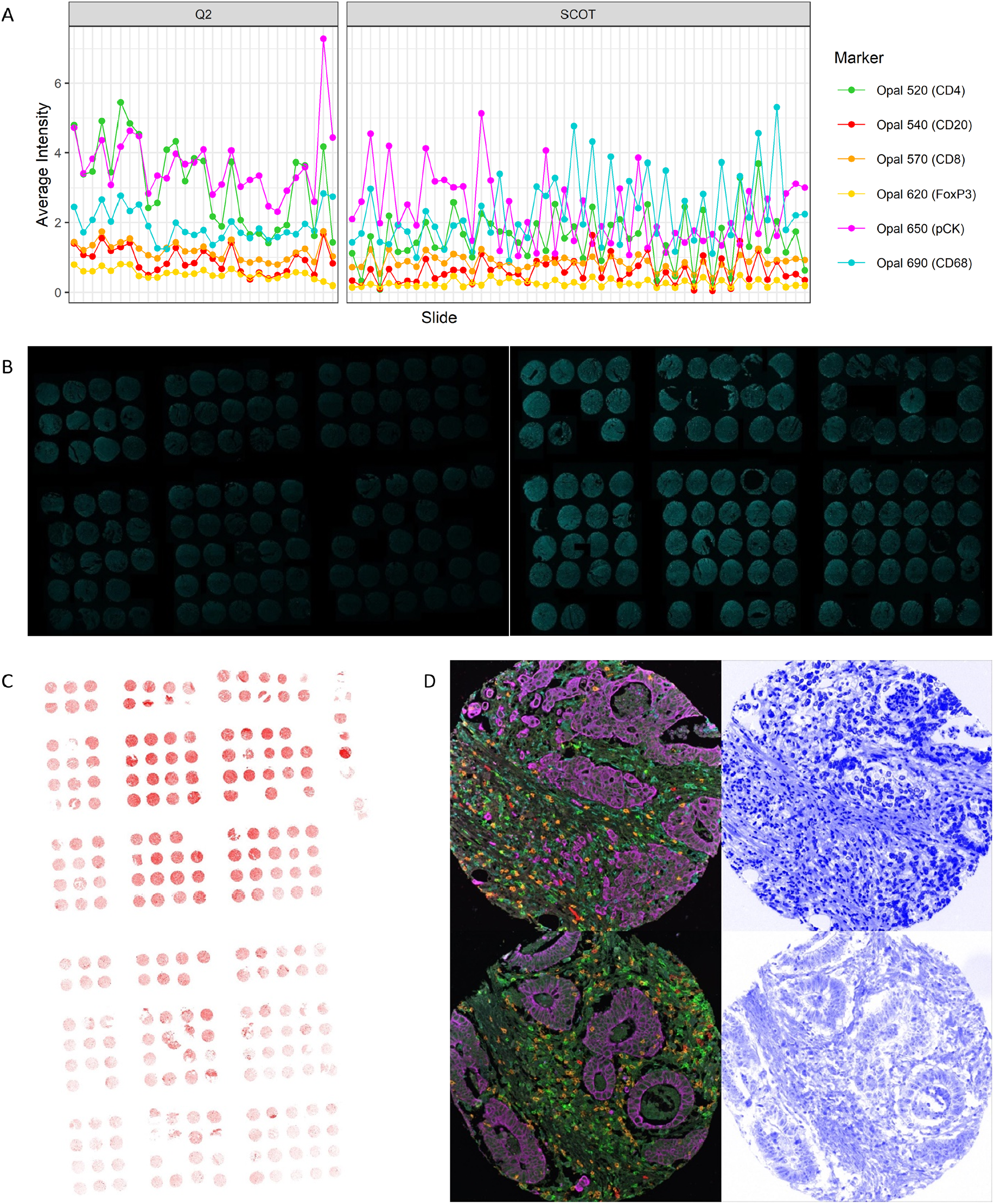
Intensity variation across the dataset. **A**. Average intensities per slide and marker across the dataset. **B**. Examples of CD8 staining (Opal 570) of different slides from the Q2 cohort illustrating inter-slide variation of single marker channels. Both images were taken with the same view settings. **C**. Example of CD20 staining (Opal 540) of a slide from the SCOT cohort illustrating intra-slide variation of single marker channels. **E**. Spot with distortion of nuclear signal (bottom row) compared to spot without distortion of nuclear signal (top row). Left: all channels except DAPI; right: DAPI channel. All images were taken with the same view settings.

### Image Analysis

The scanned TMA slides were analysed using HALO v3.4 (Indica Labs, Albuquerque, NM, USA). First, we segmented the TMA WSIs into square images of individual spots. Empty spots, spots with low amounts of tissue and spots with large staining artefacts (e.g. due to dust or air bubbles), blurry regions, tissue artefacts, tissue floaters or folds were excluded from the analysis. After exclusions, 12,592 valid spots remained in total for further analysis (for a detailed flow diagram according to REMARK guidelines [19] see **Figure 3**). We trained a deep learning algorithm for classification of the images into different regions, namely Tumour, Stroma, Muscle, Necrosis, Folds and Background using pathologist-validated tissue regions. For this purpose, we annotated a representative selection of each class and then trained the algorithm (HALO AI DenseNet V2) with these annotations. While the tissue classes Tumour and Stroma represent the classes of interest for spatially resolving marker expression analysis, the classes Necrosis, Folds, Muscle and Background were used for the exclusion of non-informative regions. The marker analysis was performed using marker-specific binary thresholds to classify cells or pixels as positive or negative, depending on whether the marker signal intensity was above or below the threshold. Pan-cytokeratin was used for tissue classification only and was not quantitatively evaluated on the cell- or pixel-level. CD4 (Opal 520) was excluded from marker analysis due to a low signal-to-noise ratio, especially in the epithelium area, where strong autofluorescence in the 500-550 nm range was observed. In a subset of slides in the SCOT cohort, we observed an increased bleed-through of the pan-cytokeratin channel (Opal 650) into the CD68 (Opal 690) channel. For these samples, intraepithelial CD68 data was excluded from further study.

**Figure 3:**
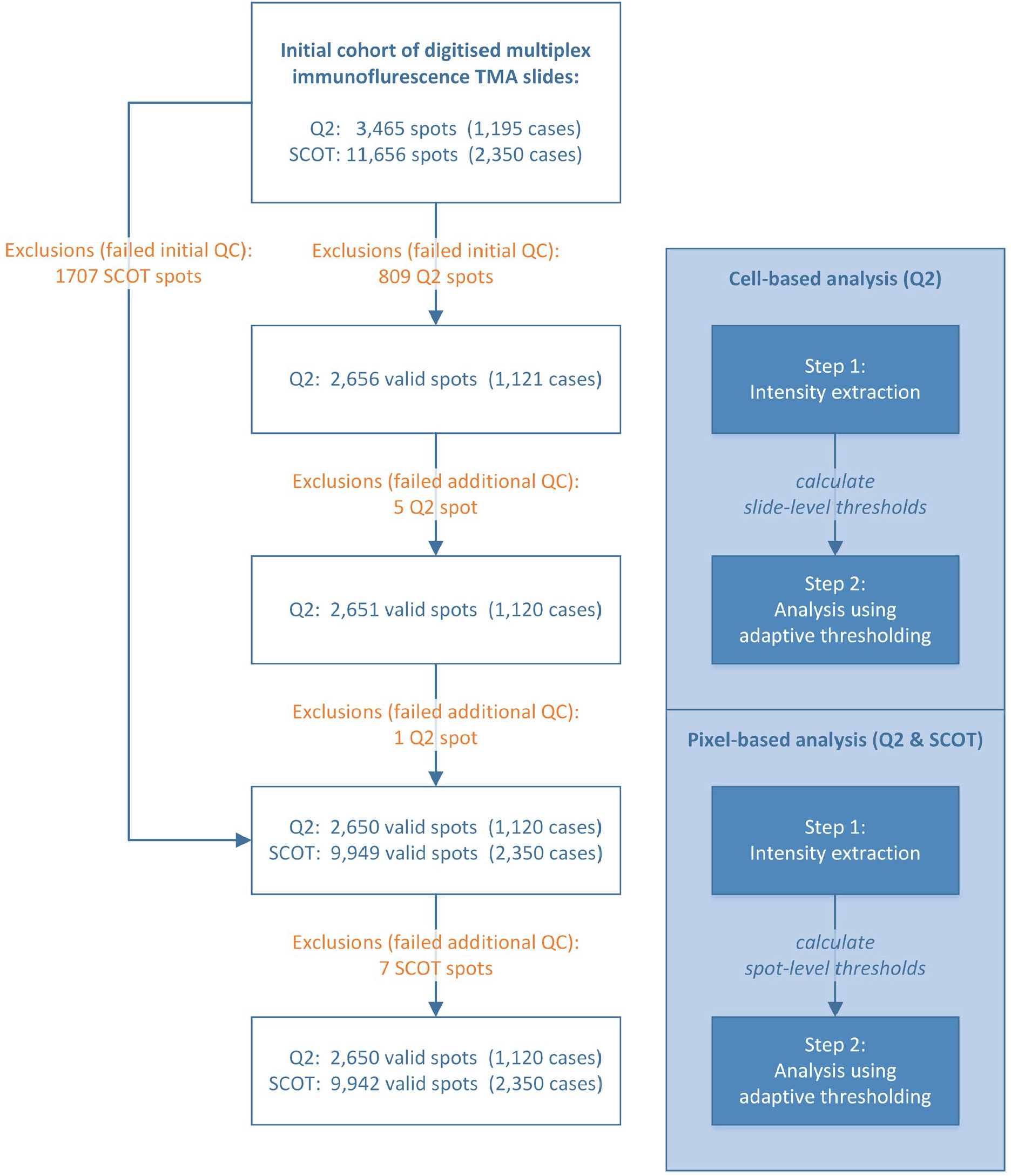
Schematic analysis workflow. Schematic visualisation of the analysis workflow and corresponding numbers of included/excluded spots and cases based on manual quality control (QC).

### Cell-based marker analysis

In the cell-based marker analysis approach, the marker expression is evaluated per cell based on the segmentation of individual nuclei using a pre-trained nuclei segmentation network from the HALO AI platform within the HALO HighPlex FL v4.0.3 analysis module. The baseline cell segmentation was refined by setting cell morphometry parameter constraints, such as nuclear size and roundness. The cytoplasm of each cell was defined as the region around the nucleus within a radius of 1 μm (or half the distance to the neighbouring cell nuclei, if the distance between two cells was lower than 1 μm). The marker expression was evaluated separately for each cell compartment (nucleus and cytoplasm). We applied the cell-based marker analysis approach to the Q2 cohort for CD8, CD20 and FoxP3 with adaptive thresholding using slide-specific marker thresholds. We tested cell-based analysis for CD68, but based on the irregular shape and large cell size of macrophage infiltrates gave preference to pixel-based analysis for CD68 from cell-based analysis in consistency with prior work [20].

### Pixel-based marker analysis

In the pixel-based marker analysis, the marker expression is not evaluated per cell or cell compartment but per pixel. A pixel was classified as marker-positive or -negative using the HALO AreaQuantification FL v2.1.10 analysis module. We applied the pixel-based marker analysis to the Q2 cohort and the SCOT cohort for CD8, CD20, CD68 and FoxP3 with adaptive thresholding using spot-specific marker thresholds.

### Statistical analysis

The image analysis results were exported from *HALO* as .csv files and analysed in *RStudio* with *R* (version 4.1.2). The correlation between cell-based and pixel-based analysis results and the correlation between mIF-derived CD8 data and IHC-derived CD8 data was assessed using Spearman’s rank correlation test and expressed by Spearman’s correlation coefficient.

## RESULTS

### Development of Adaptive Thresholding Methods for TMA cohorts

For developing an adaptive thresholding approach for application on large TMA cohorts, we extracted, separately for each marker, the average marker signal intensity for each slide in the cohort and the average marker intensity of each TMA spot. The slide-level average marker signal intensities were used to calculate the slide-specific marker thresholds. The spot-level average marker intensities were used for developing a method for calculating spot-specific marker thresholds. Due to their relatively small size, we considered individual TMA spots as sufficiently homogeneous to apply a single threshold on the entire spot. Based on the observation that global staining intensity gradients run smoothly across an entire slide, while spots with high intensity due to biological variation are distributed sparsely across the slide, we implemented a comparison with neighbouring spots to get a good estimation of the local background intensity while preserving the biologically relevant outliers. We tested different methods to aggregate the spot-level marker intensities into local marker threshold values for each single TMA spot including variation in the size of the neighbourhood that is taken into account for the calculation of the threshold of a single spot, and weighting of the influence of each spot during the calculation. The different methods were evaluated based on a systematic comparison with the ground truth defined by pathologist visual review identifying a combination of slide-specific and spot-specific thresholds as the optimal approach for TMA marker analysis to account for intra- and inter-slide intensity variation as described below. Visual assessment of the image analysis results with and without adaptive thresholding by pathologist experts showed that the analysis using adaptive thresholding achieved more accurate marker analysis and delineation of positive vs. negative pixels/cells (see **Figure 4** for visualisations).

**Figure 4:**
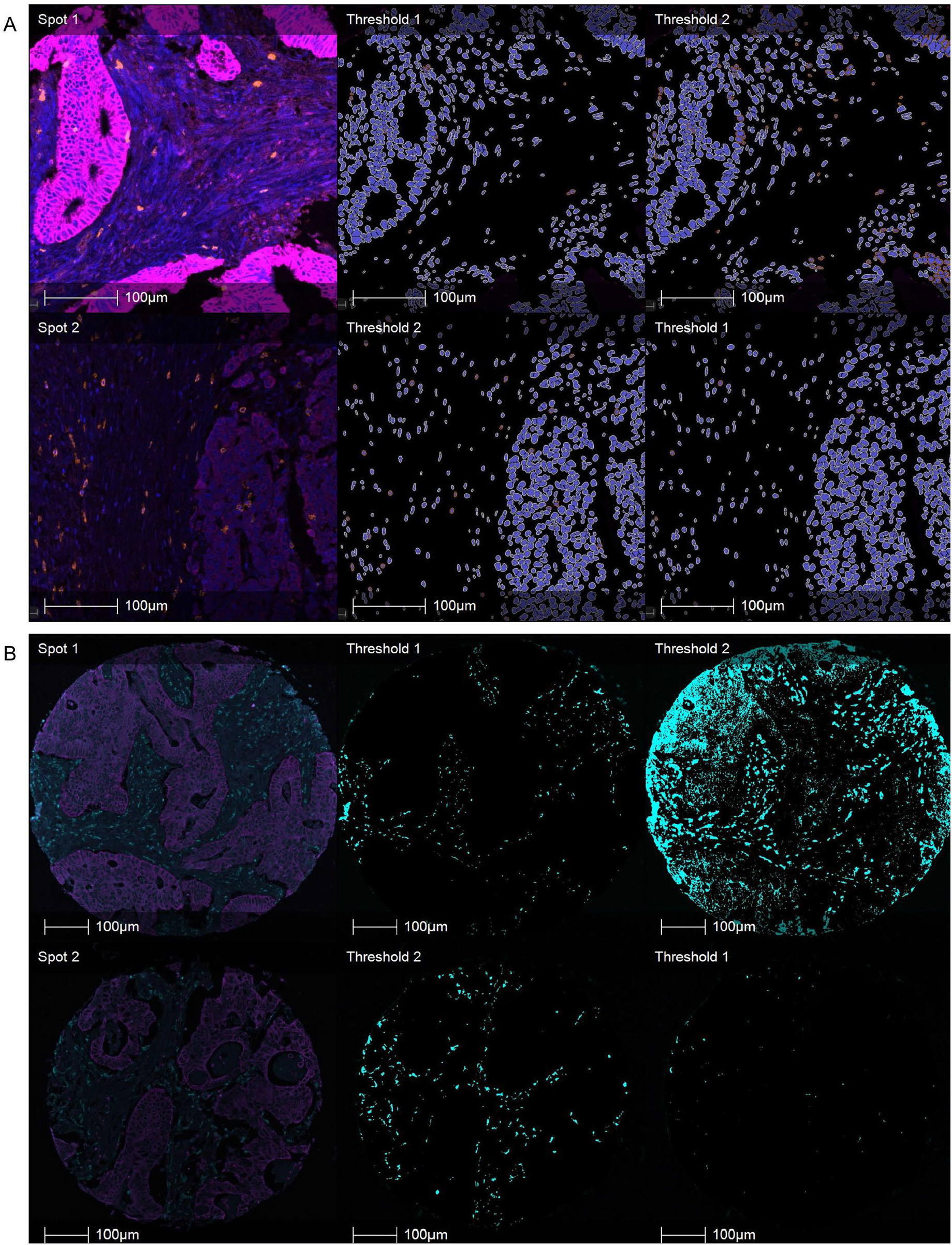
Visual comparison of image analysis with and without adaptive thresholding. **A**. Example from the Q2 cohort for cell-based marker analysis (CD8) with and without slide-specific thresholding. The two spots are sourced from different slides. Left: original image (blue: DAPI channel; orange: CD8 channel; magenta: pan-cytokeratin channel); middle: cell-level markup using suggested slide-specific threshold; right: cell-level analysis markup using slide-specific threshold suggested for the other spot, simulating global thresholding. Cells marked as marker-positive are indicated by orange cytoplasm in the analysis markup. **B**. Example from the SCOT cohort for pixel-based marker analysis (CD68) with and without spot-specific thresholding. Both spots are from the same slide. Left: original image (turquoise: CD68 channel; magenta: pan-cytokeratin channel); middle: pixel-level markup using suggested spot-specific threshold; right: pixel-level analysis markup using spot-specific threshold suggested for the other spot, simulating global thresholding. Pixels marked as CD68-positive are indicated by turquoise colour in the analysis markup.

### Accounting for Inter-Slide Variation: Calculating Slide-Specific Thresholds

For calculating slide-specific marker thresholds, we first extracted the mean marker signal intensity (mean intensity of all cells) for each slide in the cohort, separately for each marker. We defined a slide without noticeable artefacts that served as a reference and set the intensity thresholds 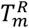 for this reference slide based on pathology review, separately for each marker *m*. After that, the marker thresholds 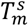 for the other slides were calculated based on the average intensity 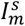 of the slide *s* compared to the average intensity 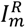 of the reference slide: 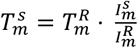

### Accounting for Intra-Slide Variation: Calculating Spot-Specific Thresholds

For calculating marker thresholds for each TMA spot individually, we first extracted the average marker intensities (average intensity of all pixels) for each channel and TMA spot location. The threshold for each spot was calculated individually based on the marker intensity of the spot itself and that of its neighbouring spots. For the final analysis, the marker threshold 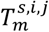 for the spot with position *i, j* in the TMA grid of slide *s* and marker *m*, was calculated as a weighted median of the intensity 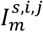 (with the amount of valid tissue 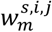 serving as corresponding weight) of the spot itself and the intensities of all spots which lie within a square of side length 7 centred on the spot, multiplied with a marker-specific factor 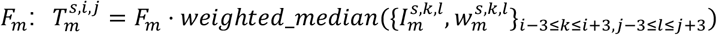. The median function was chosen due to its property to be not affected by single outliers (in contrast to the mean function, which is heavily influenced by strong outliers). In our case, when spots with very high intensity compared to neighbouring spots are observed due to biological reasons, this property was extremely helpful, since we aimed to control for the background intensity and not to smooth the overall intensity values. The amount of valid tissue in each spot served as a weight for the calculation of the weighted median. Thus, spots with greater amounts of valid tissue, which are more informative, get more weight in the calculation, and empty or invalid spots were ignored for the threshold calculation.

The marker-specific factors were set by visual assessment. The following values were set: *F*_*CD*20_ = 10, *F*_*CD*8_ = 6, *F*_*Fox*3_ = 8, *F*_*CD*68_ = 2.5. For the threshold calculation for CD68, whose analysis results are more sensitive to the choice of threshold, we added two additional features: 1) We noticed that for spots with high marker intensity, higher thresholds are more appropriate. Therefore, the weight of the centre spot (the spot for which the threshold is calculated) was multiplied by the number of neighbouring spots to give it equal weight as all the other spots together. 2) Since the spots at the boundary of the slides miss a balanced neighbourhood, we added a virtual complement for these spots, to achieve a balanced neighbourhood for all spots. For all positions where a spot is missing, we determined the marker intensity of the spot on the opposite side in the direction of the centre spot, subtracted the mean value of all present spots, and replaced the missing value with the result.

### Validation of Pixel-Based Analysis

To check the robustness of the pixel-based marker analysis, we compared the distribution of the marker densities derived from pixel-based analysis (percentage of positive area per spot) between the Q2 cohort and the SCOT cohorts (see **Figure 5A**). This comparison showed a similar distribution of the marker densities for both datasets, indicating consistency of the analysis results across both cohorts. For cross-validation of the pixel-based analysis against the cell-based analysis, we checked the correlation between the results of both analysis types. This comparison was performed on the 2,650 TMA cores of the Q2 cohort. The correlation was calculated separately for the epithelium and the stroma tissue due to their different characteristics regarding bleed-through and marker expression. The correlation between the number of positive cells and the size of the positive area (in μm^2^), were calculated by Spearman’s rank test (Spearman’s correlation coefficient ρ, see **Figure 5B)**. For CD8 and FoxP3 we determined a very strong correlation both in the stroma and the epithelium tissue (ρ ≥ 0.9). For CD20, a very strong correlation in the stroma (ρ > 0.9), and a moderate correlation in the epithelium (ρ = 0.59) was observed. In concordance with the clinical importance of the densities of positive cells per area and the percentage of positive area per total area, we also calculated the correlation between the densities of positive cells (cells per mm^2^) and the percentage of the positive area from the total area (see **Figure 5C)**. We found a very strong correlation in both tissue compartments (ρ > 0.9) for CD8. For FoxP3 and CD20, a very strong correlation in the stroma (ρ > 0.9) was seen, whereas a strong correlation for FoxP3 in the epithelium (ρ = 0.84) and a moderate correlation for CD20 in the epithelium (ρ = 0.57) was observed. For all groups, the p-value is close to 0 (p ≪ 0.01). For the exact Spearman’s correlation coefficient, we refer to **Figure 5B-C**.

**Figure 5:**
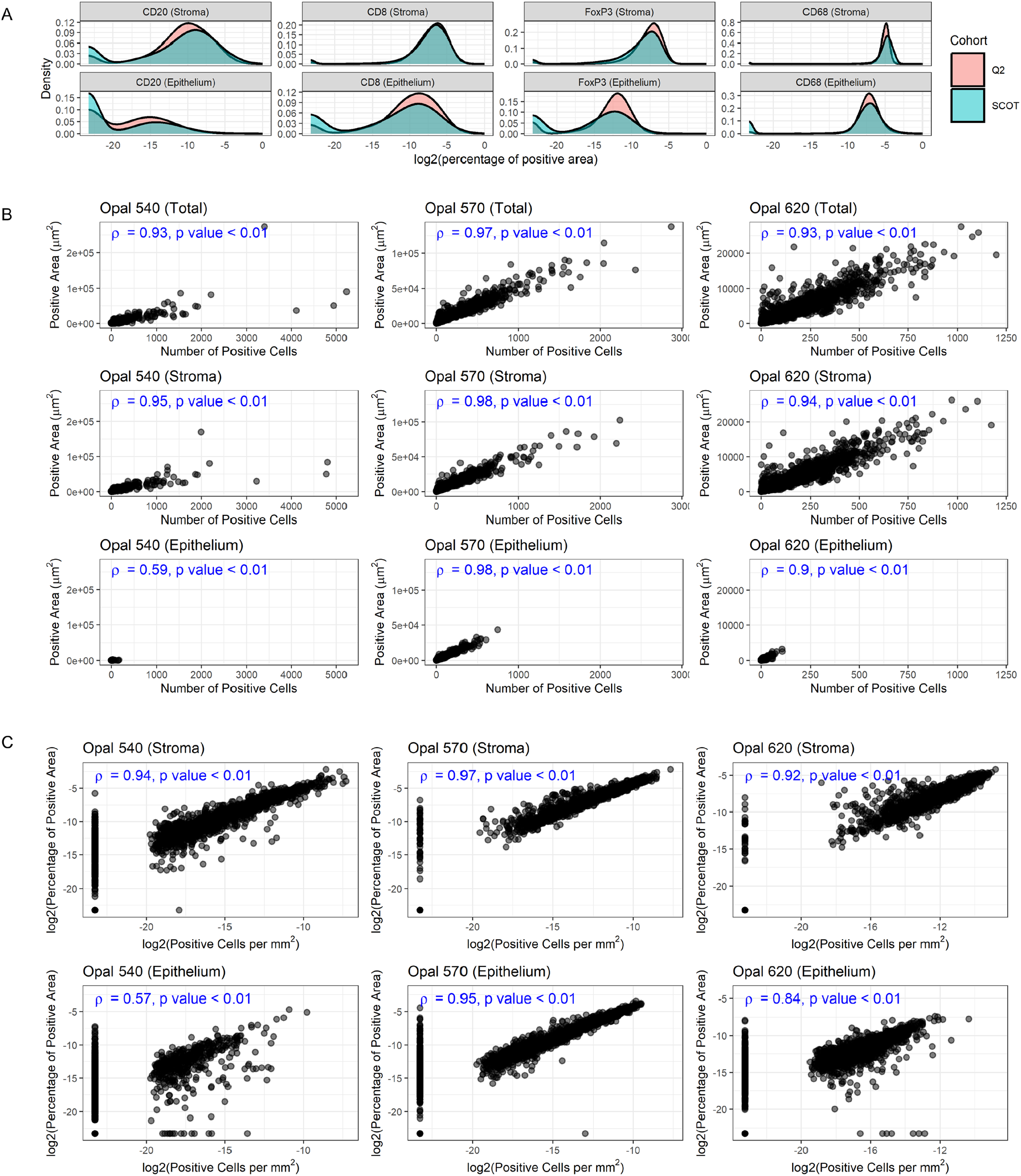
Pixel-based analysis: Density distribution and comparison with cell-based analysis. **A**. Comparison of the density distribution across the Q2 and the SCOT cohort. **B**. Comparison of absolute measurements: number of positive cells versus the amount of positive area, separated per marker and stromal/epithelial compartment, in the Q2 cohort. **C**. Comparison of density measurements: number of positive cells per area versus the amount of positive area in relation to the total area, separated per marker and stromal/epithelial compartment, in the Q2 cohort.

### Validation of Multiplex Analysis

For CD8, we compared the pixel-based analysis results in the Q2 cohort against IHC measurements from the same cohort provided by [21]. In consistency with the IHC data, we aggregated the spot-level data into case-level data by adding up spot-level cell numbers and area counts and calculated density measurements on a case level. We then compared the mIF-derived number of CD8+ cells and amount of CD8+ area across the whole tissue area with the IHC-derived number of CD8+ cells per spot and the corresponding fraction of CD8+ cells by IHC of the total number of cells or per total area, respectively. For all comparisons we see a moderate correlation (ρ between 0.63 and 0.65, with p ≪ 0.01), see **Figure 6**.

**Figure 6:**
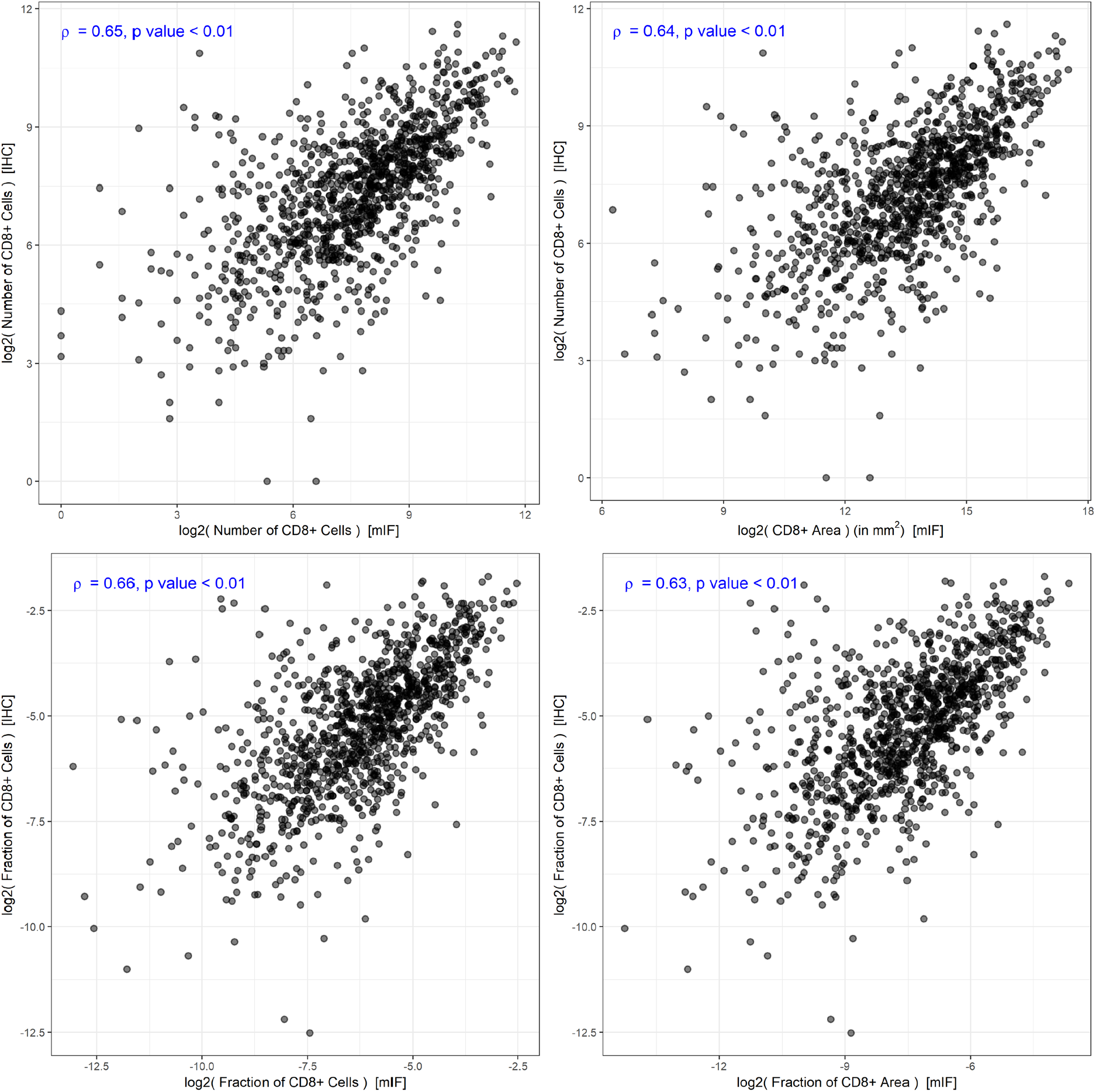
Comparison of multiplex analysis with orthogonal method in Q2 cohort. Comparison of mIF-derived with IHC-derived data about CD8-positivity. Top row: comparison of absolute measurements. Bottom row: comparison of density measurements. Left column: comparison with cell-level mIF data. Right column: comparison with pixel-level mIF data.

## DISCUSSION

Digital pathology and multiplexed staining are important tools for efficient analysis of clinical trial datasets. This is recognised by the recent consensus statement of the Society for Immunotherapy of Cancer (SITC) on best practices for multiplex IHC and IF staining and validation [13]. However, the evaluation of large cohorts with high-content image data is often compounded by a notable signal variation between as well as within images introduced by pre-analytical and analytical variables. The need for the development of standardised approaches for multiplexed IHC and IF output is equally recognised by the SITC but has not yet been addressed in guideline format. To address the unique challenges of multiplexed imaging datasets on clinical trial samples sourced from multiple institutions, we developed an adaptive thresholding method to account for both inter-slide and intra-slide variation in TMAs by digital pathology, improving the image analysis results compared to methods using a single global threshold. By comparing the results of cell-based marker analysis and pixel-based marker analysis, we show that a pixel-based marker analysis is a valid alternative to cell-based marker analysis, both when comparing the absolute number as well as density calculations of marker-positive cells. Further, by comparing the mIF image analysis results against the orthogonal IHC image analysis results, we show that the results of our image analysis are in line with established gold-standard methods and promise to confer the same prognostic impact.

This study demonstrates the value of pixel-based marker analysis in application to two large clinical trial datasets. Since a pixel-based analysis approach does not require cell segmentation, this approach enables quantitative analysis also for cell types with irregular shapes and sectioning artefacts as well as on images with insufficient nuclear signal, either intentionally left out or corrupted by error. Malesci et al. applied a pixel-based analysis approach for quantification of macrophage density and showed that a high density of macrophages in the tumour microenvironment was significantly associated with better prognosis in patients treated with 5-fluorouracil adjuvant therapy [20]. We show that there is a strong rank correlation between the results of the pixel-based analysis and the cell-based analysis, both with respect to absolute measurements as well as density measurements. The moderate correlation of CD20 in the epithelium might be affected by the very low CD20+ values in this tissue compartment where even small deviations between both measurements lead to numerically stronger effects in the correlation measurement. Taken together, these results indicate that pixel-based analysis is a valid approach for mIF fluorescence-stained slides even in the setting of moderate to large pre-analytical variation. Thus, we provide a solid reason for applying pixel-based analysis for marker analysis and provide evidence that these results can be directly compared with studies based on cell-based analysis.

Normalisation and adaptive thresholding are two closely related concepts, i.e. global thresholding on a normalised dataset can be also carried out as adaptive thresholding on non-normalised dataset. In the present study, we introduce and validate different approaches for adaptive thresholding. To account for inter-slide variation in cohorts stained by multiplexed IF imaging, different approaches are previously described. Raza et al. [22] presented a method for pre-processing normalisation of multiplexed fluorescence images using linear min-max-normalisation after noise filtering. Chang et al. [23] proposed a method accounting for inter-slide variation in multiplexed IF images, which is based on the definition of mutually exclusive markers. Thereby, they derived a set of cells which are assumed to be negative and serve as the basis to derive the background intensity level. Harris et al. [24] tested different data transformation and normalisation methods for accounting for inter-slide variation in multiplexed IF images. They found that for inter-slide variation, a division by the mean of the slide is the most accurate normalisation method while maintaining biological signals. This is in line with our method for accounting for inter-slide variation. However, none of these methods account for intra-slide variation in large cohorts, which not only consists of marker intensity variation between different images but also notable intensity variation within each slide. In this setting, it is not sufficient to compensate for inter-slide variation, but additional consideration of intra-slide variation is required.

To the best of our knowledge, no adaptive thresholding approach exists to address the considerable intra-slide variation in mIF TMA datasets including samples from a multitude of patients, institutions and regions. In the present work, we consider the image of each spot as a single image and develop an approach which accounts for inter-image variation specifically for the TMA spot images. Previously reported local thresholding methods mostly work on pixel-level and take into account the mean, median, minimal, maximal, and/or standard deviation value of a local neighbourhood of pixels for deciding whether a given pixel is considered as negative or positive [25]. Due to the nature of TMA slides, pixel-level adaptive thresholding methods are not suitable for application in this use case: If the chosen neighbourhood size is too small (smaller than the spot diameter), the background intensity gradient running across the whole slide is not captured. If the chosen neighbourhood size is larger than the spot diameter, the resulting data can be skewed by the large background area typical for TMA slides. Our proposed method accounts for inter- and intra-slide variation and solves the drawback of pixel-level adaptive thresholding methods by only taking into account tissue regions for the calculation of the spot-level thresholds. The method could be considered as an adapted median local thresholding approach applied to the TMA spots by considering each TMA spot as a single data point.

Previous reports present analyses of retrospective population-based mIF CRC TMA cohorts with 927 and 746 included patients respectively [26,27]. We are not aware of any other multiplex CRC clinical trial cohorts of comparable size or complexity in terms of the number of patients included, participating institutions or regional variation as in the present study. The analysis approach using digital pathology methods in combination with automated marker quantification allowed immunoprofiling in a high-throughput manner. Further, the availability of orthogonal data by the current gold-standard single marker IHC allowed direct cross-validation of our proposed image analysis approach, which is an additional strength of our study. As quantitative cell-based image analysis allows to link cellular identity (e.g., lineage marker expression) to a defined x-y location on the slide, future efforts could focus on determining the precise spatial relationship of specific immune cells to cancer cells in their immediate proximity (e.g., by nearest neighbour analysis) in the context of clinical outcomes.

However, our study has also some limitations. The applicability of our adaptive thresholding methods to other multiplex cohorts will have to be further tested and validated for other markers with different expression patterns. The intra-slide adaptive thresholding method is based on individual spot images and therefore not a priori applicable to non-TMA WSIs. While the cell-based analysis approach allows capturing concurrent marker-positivity for each cell individually, the pixel-based analysis approach did not allow us to capture multi-positive pixels, thus limiting the applicability to settings where the accurate quantification of well-defined lineage or functional markers is of central interest. Further technical optimisation of staining and imaging protocol may be addressed in the future to increase signal-to-noise ratio and reduce observed bleed-through artefacts, hereby enabling extended marker analysis.

In conclusion, pixel-based analysis and adaptive thresholding methods enable a reliable analysis of multiplex image cohorts showing large pre-analytical heterogeneity. Since this allows extraction of valuable information from images with pre-analytical signal heterogeneity and out-of distribution properties, this promises a broader application of digital image analysis in clinical trial datasets and facilitates the integration with clinical data. Our proposed adaptive thresholding approach accounts for variation within TMA slides and offers a method for analysing TMA images across large cohorts with considerable signal intensity variation between and within slides. Further, we provide evidence that pixel based approaches have increased robustness for the quantification of challenging marker sets or technical settings while the quantitative results remain robustly comparable to the current gold-standard approach of cell-level segmentation and quantification.

## Data Availability

The datasets pertaining to the SCOT trial used during the current study are available from the TransSCOT collaboration on reasonable request. Applications for analysis of TransSCOT samples are welcome and should be addressed to JH. Datasets and samples from the QUASAR2 trial are available upon reasonable request and should be addressed to DNC

## ADDITIONAL INFORMATION

## Acknowledgements

We would like to thank the patients who participated in the SCOT and QUASAR 2 trials and consented for their samples to be used for correlative research, as well as the recruiting clinicians and study team. We are also grateful to the Translational Histopathology Laboratory, Department of Oncology, University of Oxford, for performing immunostaining and Glasgow Tissue Research Facility, University of Glasgow, for TMA construction and scanning.

## TransSCOT consortium

The TransSCOT Trial Management Group includes (alphabetical order): David Church^1^, Enric Domingo^2^, Joanne Edwards^3^, Bengt Glimelius^4^, Ismail Gogenur^5^, Andrea Harkin^6^, Jennifer Hay^7^, Timothy Iveson^8^, Emma Jaeger^2^, Caroline Kelly^6^, Rachel Kerr^2^, Noori Maka^7^, Hannah Morgan^7^, Karin Oien^7^, Clare Orange^9^, Claire Palles^10^, Campbell Roxburgh^3^, Owen Sansom^11^, Mark Saunders^12^, Ian Tomlinson^2^.

^1^ Cancer Genomics and Immunology Group, The Wellcome Centre for Human Genetics, University of Oxford, Oxford, UK; ^2^ Department of Oncology, University of Oxford, Oxford, UK; ^3^ School of Cancer Sciences, University of Glasgow, Glasgow, UK; ^4^ Uppsala University, Uppsala, Sweden; ^5^ Centre for Surgical Science, Zealand University Hospital, Denmark; ^6^ CRUK Glasgow Clinical Trials Unit, University of Glasgow, Glasgow, UK; ^7^ Glasgow Tissue Research Facility, University of Glasgow, Queen Elizabeth University Hospital, Glasgow, UK; ^8^ University of Southampton, Southampton, UK; ^9^ NHS Greater Glasgow and Clyde Biorepository, Glasgow, UK; ^10^ University of Birmingham, Birmingham, UK; ^11^ CRUK Beatson Institute, Glasgow, UK; ^12^ The Christie NHS Foundation Trust, Manchester, UK

## Authors’ contributions

Conceptualisation: ALF, DNC, VHK

Data curation: ALF, AM, RRAKS, MAG, FJ, LG, TT, AH, TJI, MS, KO, NM, FP, LC, JH, WK, RSK, DJK, HED, ED, DNC

Formal Analysis: ALF, AM, RRAKS, FJ, LG, TT, AH, ED, DNC, VHK

Funding acquisition: OS, IT, DNC, VHK

Investigation: ALF, FJ, DNC, VHK

Methodology: ALF, DNC, VHK

Project administration: JH, DNC, VHK

Resources: ALF, MAG, AH, TJI, MS, KO, NM, FP, LC, JH, JE, OS, CK, IT, WK, RSK, DJK, HED, ED, TransSCOT consortium, DNC, VHK

Software: ALF, VHK

Supervision: DNC, VHK

Validation: ALF, DNC, VHK

Visualisation: ALF, AM, FJ, ED, DNC, VHK

Writing – original draft: ALF, VHK

Writing – review & editing: All authors

## List of abbreviations

CRC: colorectal cancer
IF: immunofluorescence
IHC: immunohistochemistry
mIF: multiplex immunofluorescence
QC: quality control
Q2: QUASAR 2
SITC: Society for Immunotherapy of Cancer
TMA: tissue microarray
WSI: whole slide image

## Ethics approval and consent to participate

Ethical approval for patient recruitment and sample collection in the SCOT and QUASAR 2 trial was obtained centrally and at all recruiting centres (REC reference number 07/S0703/136 and 04/MRE/11/18). Ethical approval for anonymised tumour molecular analysis was granted by Oxfordshire Research Ethics Committee B (REC 05\Q1605\66).

